# Efficacy of pulmonary rehabilitation in severe and critical-ill COVID-19 patients: a controlled study

**DOI:** 10.1101/2020.12.08.20245936

**Authors:** Gilbert Büsching, Zhongxing Zhang, Jean Paul Schmid, Thomas Sigrist, Ramin Khatami

## Abstract

Severe and critical ill COVID-19 patients frequently need acute care hospitalization including mechanical ventilation at ICU due to acute respiratory distress. A high proportion of these patients will develop ICU-acquired weakness and a need for rehabilitation. However data on rehabilitation outcomes in these patients are scarce and the efficacy of rehabilitation remains essentially unclear. We therefore compared the rehabilitation outcomes between COVID-19 patients with pneumonia and other patients with common pneumonia to assess their rehabilitation efficacies.

We retrospectively compared the performances of six-min walk test (6MWT), chronic respiratory questionnaire (CRQ), and functional independence measure (FIM) at the discharge from pulmonary rehabilitation between 51 Covid-19 patients and 51 patients with common pneumonia using linear regression controlled for baseline values at entrance, age, sex and cumulative Illness rating scale. Fisher exact test was applied to test whether the odd ratios (ORs) of non-improvement/improvement in 6MWT (>30-m) and CRQ (>10-point) at discharge were different between the two groups.

Covid-19 patients had similar performances at discharge in 6MWT (P-value=0.14), CRQ (P-value=0.55), and 4.2-point higher (P-value=0.004) in FIM compared to the control group. No differences in the outcomes were found between severe and critical COVID-19 patients. The OR of non-improvement/improvement in 6MWT was 0.30 (P-value=0.13) between COVID-19 and control groups; but the odd of non-improvement in CRQ tended to be 3.02 times higher (P-value =0.075) in COVID-19 group.

In-house rehabilitation is effective and suitable for COVID-19 patients irrespective from disease severity. The discrepancy of high physical improvement and relatively low gains of disease related quality of life compared to control patients with common pneumonia is however remarkable. Further studies need to evaluate whether this discrepancy is an indicator of chronic disease development.

## Introduction

Severe SARS-CoV-2 infection can lead to hospitalization due to COVID-19 pneumonia. A considerable proportion of these patients need mechanical ventilation at ICU due to acute respiratory distress and will develop ICU-acquired weakness [1, 2]. Recently increasing evidences suggest that COVID-19 patients after hospitalization may still have abnormal pulmonary function [3], low physical functioning and impaired performances of daily life activities [4], and persistent fatigue [5]. So there are increasing rehabilitation needs for COVID-19 patients who continue to suffer from the long-term consequences of COVID-19. Therefore, European Respiratory Society and American Thoracic Society-coordinated International Task Force published an interim guidance on rehabilitation for COVID-19 patients in the hospital and post-hospital [6]. However this guidance is not based on data but reflect experts’ recommendations due to the lack of data on the safety and efficacy of rehabilitation programs. Since the number of patients with SARS-CoV-2 infection is exponentially increasing, both, the patients and healthcare professionals cannot wait for published randomized controlled trials before starting the rehabilitative interventions. So far, only one pilot study reported that current pulmonary rehabilitation used for pneumonia patients can improve physical performance in six-in walk test (6MWT) of critical COVID-19 patients who were admitted to ICU [7]. In this study, the efficacy of rehabilitation outcome remained unclear for two reasons. First, the outcome data were not compared to a control group. Second, assessment of the minimal clinically important difference (MCID) [8, 9] is missing making it difficult to estimate the relevance of clinical improvement. Thus, it remains unclear whether rehabilitation programs applied to COVID-19 patients can achieve similar improvements in the physical functioning and quality of life as compared to other pneumonia patients.

We therefore investigated the clinical course of COVID19 patients during rehabilitation and compared the outcomes of physical performance (i.e., 6MWT), functional independence measure (FIM) [10] and chronic respiratory questionnaire (CRQ) [11, 12] to matched cases suffering from other types of pneumonia who attended the same rehabilitation program. We were specifically interested in the proportion of patients who failed to achieve a clinically relevant improvement as indexed by the MCID and the impact of ICU-treatment for their outcome.

## Method

We retrospectively investigated a cohort of 51 COVID-19 patients with pneumonia referred from acute care hospitals to inpatient pulmonary rehabilitation at Department of Pulmonology, Clinic Barmelweid AG from March 23^rd^ to May 29^th^ 2020. We compared these COVID-19 patients with 51 consecutive pneumonia patients (“common pneumonia”), who did the same rehabilitation protocol in 2019 in our clinic. Patients received standard pulmonary rehabilitation [13], defined after the national requirements from medical faculty [14]. The common pneumonia patients in the control group were selected according to following criteria: 1) older than 40 years; 2) no history of thoracic and/ or pulmonary surgery; 3) complete set of outcome data; and 4) did not repeat the rehabilitation program in 2019. All Patients gave written general consent for using their data for research purpose. The patients’ data were anonymised before analysis. This study was approved by the scientific committee of Academy Barmelweid.

Physical performance (6MWT), CRQ, and FIM were measured at the entrance and at discharge from the rehabilitation program (see Table 1). The mean values (IQR) of COVID-19 patients were compared to the ones of the common pneumonia control group both at entrance baseline and at discharge. The improvements in 6MWT, CRQ and FIM calculated as the differences between discharge and the baseline were compared between the two groups. The numbers of patients whose improvements reached the established MCID: 30-m for 6 MWT [15] and 10-point for CRQ were also calculated in the two groups. We chose 10-point for CRQ because we used the original score of CRQ which has a full score of 140 points. This 10-point equalled to the 0.5-point MCID if it was converted to the 0-7 points system [16].

**Table 1:**
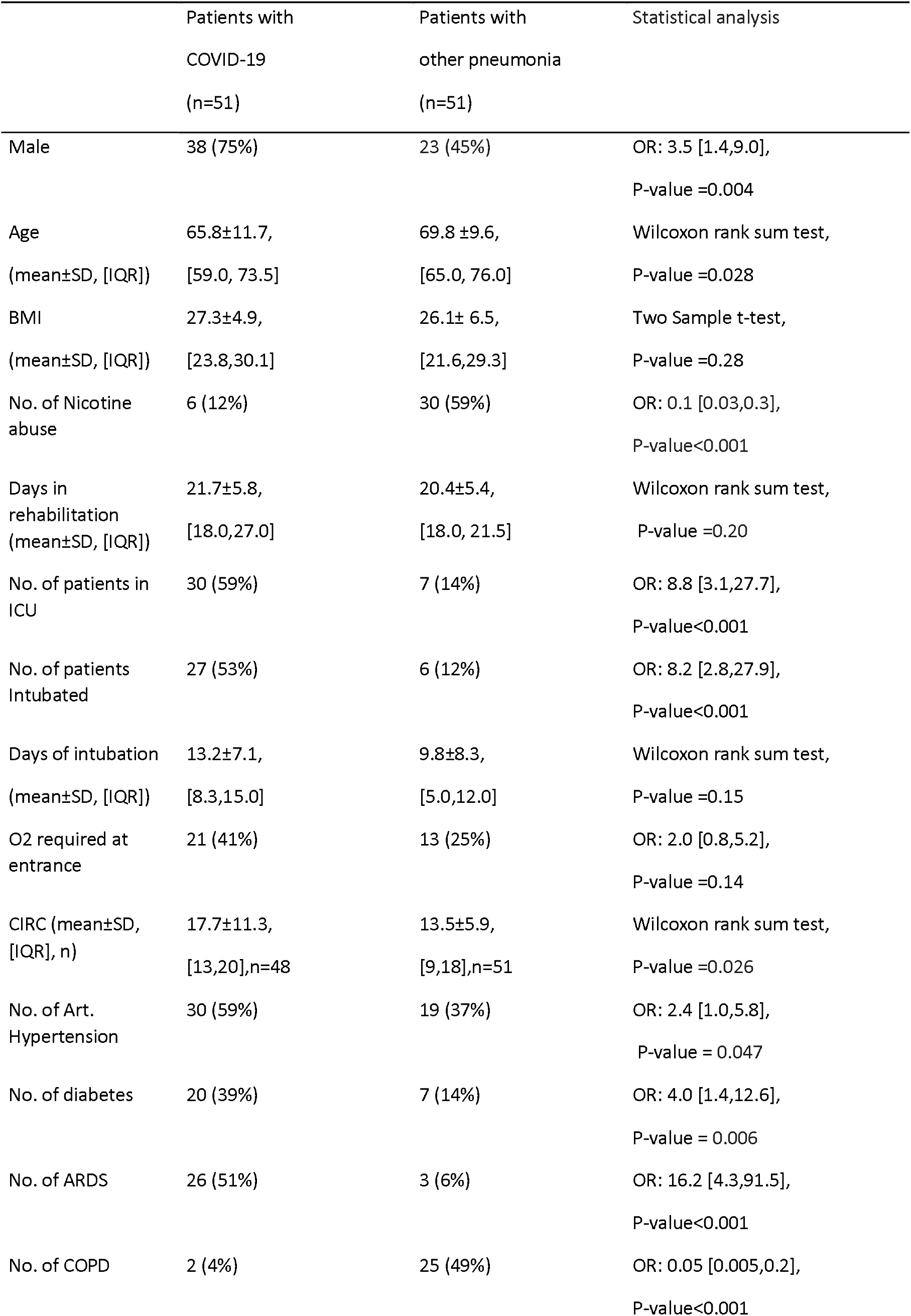
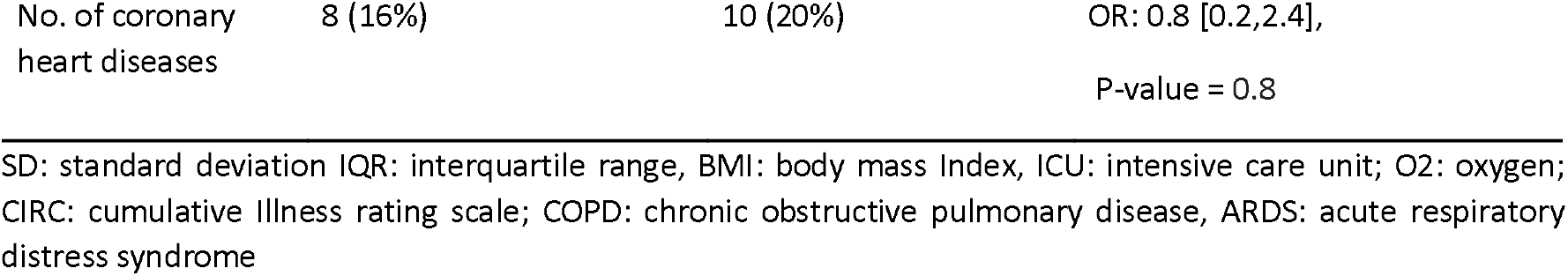
Patient demography

|  | Patients with<br>COVID-19<br>(n=51) | Patients with<br>other pneumonia<br>(n=51) | Statistical analysis |
| --- | --- | --- | --- |
| Male | 38 (75%) | 23 (45%) | OR: 3.5 [1.4,9.0],<br>P-value =0.004 |
| Age<br>(mean±SD, [IQR]) | 65.8±11.7,<br>[59.0, 73.5] | 69.8 ±9.6,<br>[65.0, 76.0] | Wilcoxon rank sum test,<br>P-value =0.028 |
| BMI<br>(mean±SD, [IQR]) | 27.3±4.9,<br>[23.8,30.1] | 26.1± 6.5,<br>[21.6,29.3] | Two Sample t-test,<br>P-value =0.28 |
| No. of Nicotine<br>abuse | 6 (12%) | 30 (59%) | OR: 0.1 [0.03,0.3],<br>P-value<0.001 |
| Days in<br>rehabilitation<br>(mean±SD, [IQR]) | 21.7±5.8,<br>[18.0,27.0] | 20.4±5.4,<br>[18.0, 21.5] | Wilcoxon rank sum test,<br>P-value =0.20 |
| No. of patients in<br>ICU | 30 (59%) | 7 (14%) | OR: 8.8 [3.1,27.7],<br>P-value<0.001 |
| No. of patients<br>Intubated | 27 (53%) | 6 (12%) | OR: 8.2 [2.8,27.9],<br>P-value<0.001 |
| Days of intubation<br>(mean±SD, [IQR]) | 13.2±7.1,<br>[8.3,15.0] | 9.8±8.3,<br>[5.0,12.0] | Wilcoxon rank sum test,<br>P-value =0.15 |
| O2 required at<br>entrance | 21 (41%) | 13 (25%) | OR: 2.0 [0.8,5.2],<br>P-value =0.14 |
| CIRC (mean±SD,<br>[IQR], n) | 17.7±11.3,<br>[13,20],n=48 | 13.5±5.9,<br>[9,18],n=51 | Wilcoxon rank sum test,<br>P-value =0.026 |
| No. of Art.<br>Hypertension | 30 (59%) | 19 (37%) | OR: 2.4 [1.0,5.8],<br>P-value = 0.047 |
| No. of diabetes | 20 (39%) | 7 (14%) | OR: 4.0 [1.4,12.6],<br>P-value = 0.006 |
| No. of ARDS | 26 (51%) | 3 (6%) | OR: 16.2 [4.3,91.5],<br>P-value<0.001 |
| No. of COPD | 2 (4%) | 25 (49%) | OR: 0.05 [0.005,0.2],<br>P-value<0.001 |

SD: standard deviation |QR: interquartile range, BMI: body mass Index, ICU: intensive care unit; O2: oxygen; CIRC: cumulative illness rating scale; COPD: chronic obstructive pulmonary disease, ARDS: acute respiratory distress syndrome

Odd ratio (OR) and its 95% confidence interval (CI) were calculated to compare the binary parameter between the control group and the COVID-19 group (Fisher’s exact test). Non-parametric Wilcoxon rank sum test was used to compare the numerical variables between the two groups if their distributions were not normal. The normality was tested by Shapiro-Wilk test. For numerical variable with normal distribution, we first compared its variances between the two groups (F-test). Then two sample t-test or Welch two sample t-test was applied if it had equal or unequal variance between the two groups, respectively. Fisher’s exact test was applied to test whether the OR of non-improvement (i.e., failed in MCID)/improvement (i.e., achieved MCID) in 6MWT and CRQ at discharge were different between the two groups. The significance level was P-value<0.05.

Linear regressions were used to compare whether the 6MWT distance, FIM and CRQ scores were different between the two groups at discharge, after controlling their values at entrance baseline, age, sex and cumulative Illness rating scale (CIRS). We further tested whether the outcomes of our rehabilitation program were different between severe and the critical patients (i.e., non-ICU vs. ICU) using linear regression after controlling the aforementioned variables. This subgroup analysis can test whether the similar efficacy of the rehabilitation outcome can be also achieved in critical COVID-19 patients because many COVID-19 patients were admitted to ICU during hospitalization and these patients are more likely to have higher and longer-term healthcare/rehabilitation burden. All statistical analyses were done using R (version 3.2.4).

## Results

### Patient demography

Patients’ demography at the entrance is shown in Table 1. Most of our COVID-19 patients were male and younger than controls. They had higher CIRS scores at entrance, probably because more COVID-19 patients were treated at ICU and had mechanical ventilation compared to control group. COVID-19 patients had higher odd to have the comorbidities of hypertension, diabetes and acute respiratory distress syndrome (ARDS), while the control group had higher odd of having comorbid chronic obstructive pulmonary disease (COPD).

### Outcome of rehabilitation: COVID-19 patients vs. common pneumonia patients

At baseline the two groups had similar performance in 6MWT while COVID-19 patients had better CRQ and FIM scores (Table 2). The changes in the performances of 6MWT, CRQ and FIM compared to baseline in each patient are shown in Fig.1, indicating that the majority of patients improved in both groups. Paired t-tests confirmed that both groups had significantly improvements in 6MWT, in CRQ, and in FIM (Table 2). At discharge COVID-19 patients achieved better performances in 6MWT and FIM but similar CRQ scores after our rehabilitation program compared to control group (Table 2). Likewise COVID-19 patients tended to have higher degree of improvement in 6MWT compared to the control group, whereas the increase in CRQ and FIM were similar in both groups with no significant differences (Table 2).

**Figure 1.**
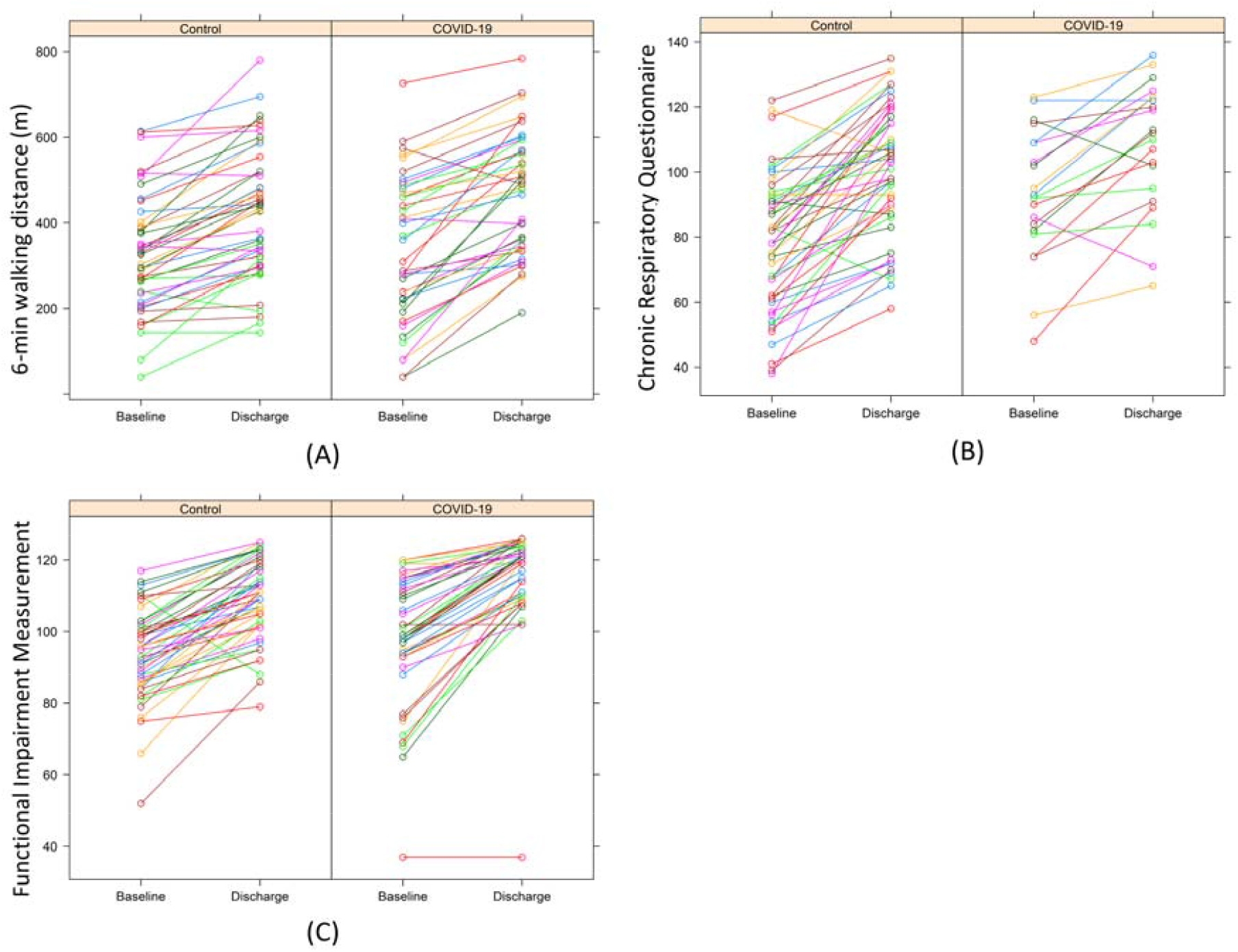
The performances of 6-min walking test (A), chronic respiratory questionnaire (B) and functional impairment measurement (C) in each patient at the baseline (i.e., entrance) and at the discharge of our pulmonary rehabilitation. In general the performance improves in both control group (left) and the COVID-19 group (right).

**Table 2:**
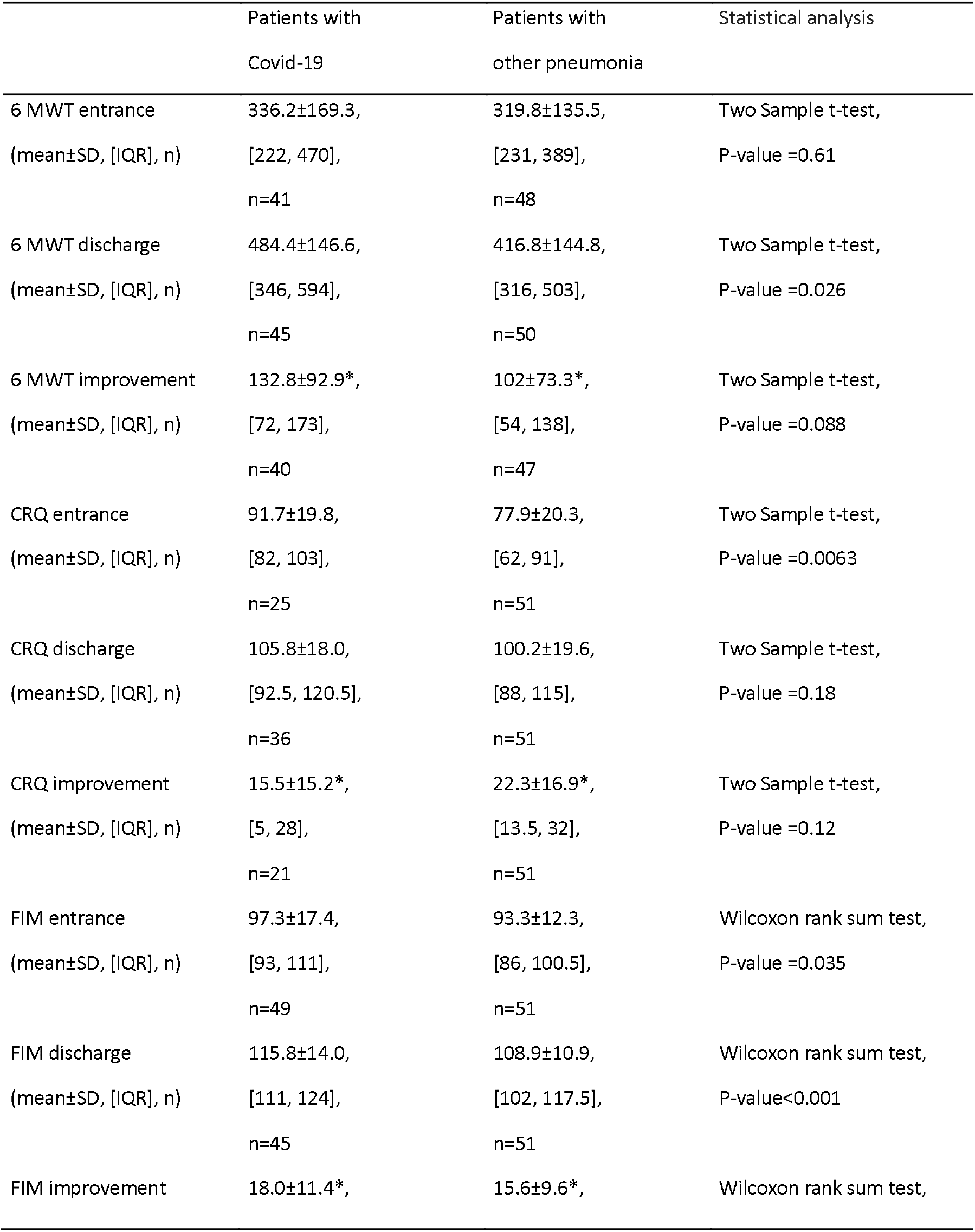
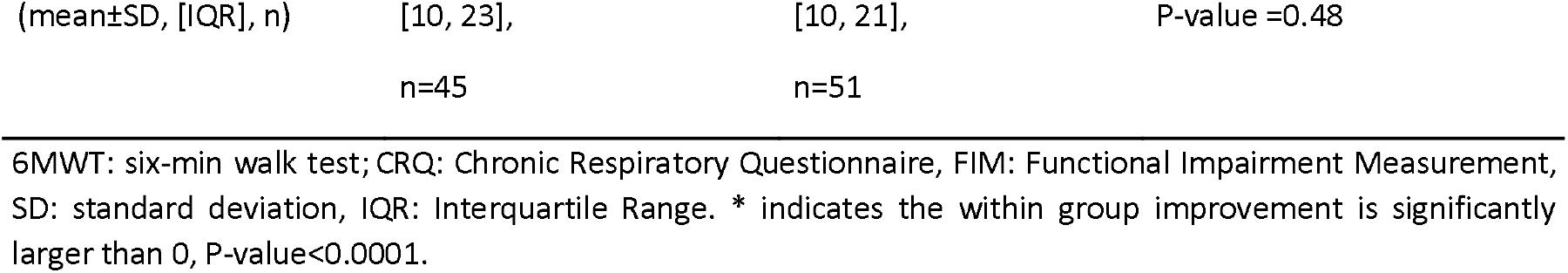
The comparisons of 6MWT, CRQ and FIM between the two groups

Considering the differences in age, sex and CIRS scales at baseline between the two groups, we performed a regression analysis to test whether the outcomes at discharge were still significantly different after controlling for these variables and for the baseline performances. The results are shown in Table 3. Data of 37 COVID-19 and 47 control patients were used to build the regression model of 6MWT after deleting patients with missing values and outliers defined by nonparametric boxplot. The numbers of patients were 21 and 51 for the model of CRQ, 44 and 49 for the model of FIM, respectively. There were no significant differences in the 6MWT performance and CRQ at discharge between these two groups, but COVID-19 patients had on average 4.16 points higher (P-value=0.00364) in FIM score compared to control group.

**Table 3.** The results of regression analysis of COVID-19 group vs. control group

|  | 6MWT at discharge<br>(Adjusted R<sup>2</sup>=0.75) |  |  | CRQ at discharge<br>(Adjusted R<sup>2</sup>=0.75) |  |  | FIM at discharge<br>(Adjusted R<sup>2</sup>=0.75) |  |  |
| --- | --- | --- | --- | --- | --- | --- | --- | --- | --- |
|  | Estimate | Std.<br>Error | P-value | Estimate | Std.<br>Error | P-value | Estimate | Std.<br>Error | P-value |
| Age | -2.06 | 0.81 | 0.0134 | -0.23 | 0.18 | 0.202 | -0.05 | 0.06 | 0.432 |
| Sex:m | 34.11 | 17.56 | 0.0557 | 1.33 | 3.77 | 0.726 | 2.45 | 1.35 | 0.072 |
| 6MWT Baseline | 0.70 | 0.06 | <0.0001 | / | / | / | / | / | / |
| CRQ Baseline | / | / | / | 0.62 | 0.09 | <0.0001 | / | / | / |
| FIM Baseline | / | / | / | / | / | / | 0.45 | 0.05 | <0.0001 |
| CIRC | -1.64 | 0.92 | 0.0787 | -0.42 | 0.62 | 0.184 | 0.005 | 0.075 | 0.942 |
| COVID-19 | 26.55 | 17.61 | 0.136 | -2.70 | 4.45 | 0.545 | 4.16 | 1.39 | 0.00364 |

6MWT: six-min walk test; CRQ: Chronic Respiratory Questionnaire, FIM: Functional Impairment Measurement; CIRC: cumulative illness rating scale.

Next we compared the number of patients who did not achieve clinically relevant outcome according to MCID. A proportion of 7.5% (3/40) COVID-19 and 21.3% (10/47) control patients failed to meet MCID of 6MWT after the rehabilitation program. Fisher’s exact test showed that the OR of non-improvement/improvement between the COVID-19 and control groups was 0.30 (95% CI: 0.05-1.31, P-value=0.13). So the odd of no improvement in 6MWT in COVID-19 group was not different from the one of the control group, although the proportion appeared to be smaller in the COVID-19 group (i.e., 7.5% vs. 21.3%).

An even higher proportion of 42.9% (9/21) COVID-19 patients did not reach MCID of CRQ after the rehabilitation program. This number was 19.6% (10/51) in the control group, resulting in an OR of 3.02 (95% CI: 0.87-10.64, P-value =0.075) of non-improvement/improvement in CRQ between the COVID-19 and control groups, i.e., the odd of no effect in improving CRQ showed a trend to be 3.02 times higher in COVID-19 group compared to the control group. In the nine COVID-19 patients who failed in MCID six patients were critical ill at ICU and intubated for mechanical ventilation, six patients were male and four patients were without ARDS. These smaller numbers of patients limited us to further explore which factors may predict the failure of improving CRQ via regression analysis.

### Outcome of rehabilitation: COVID-19 patients ICU vs. non-ICU

Data of 23 critical COVID-19 patients who were treated at ICU and 14 severe COVID-19 patients who were not at ICU were used to build the regression model of 6MWT after deleting patients with missing values and outliers. The dependent variable was the 6MWT at discharge, and age, sex and CIRC scales and baseline 6MWT performance were controlled in the model. The results are shown in Table 4 below. There was no significant difference in the 6MWT at discharge between the two subgroups, indicating that the physical functioning outcome of our rehabilitation program is similar between critical and severe COVID-19 patients.

**Table 4.** The results of regression analysis of COVID-19 ICU subgroup vs. non-ICU subgroup

|  | 6MWT at discharge<br>(Adjusted R<sup>2</sup>=0.77) |  |  | CRQ at discharge<br>(Adjusted R<sup>2</sup>=0.40) |  |  | FIM at discharge<br>(Adjusted R<sup>2</sup>=0.61) |  |  |
| --- | --- | --- | --- | --- | --- | --- | --- | --- | --- |
|  | Estimate | Std. Error | P-value | Estimate | Std. Error | P-value | Estimate | Std. Error | P-value |
| Age | -2.37 | 1.09 | 0.0379 | -0.11 | 0.31 | 0.722 | -0.11 | 0.069 | 0.11 |
| Sex:m | 55.38 | 26.77 | 0.047 | 8.91 | 7.96 | 0.280 | 3.71 | 1.62 | 0.027 |
| 6MWT Baseline | 0.57 | 0.073 | <0.0001 | / | / | / | / | / | / |
| CRQ Baseline | / | / | / | 0.68 | 0.18 | 0.0021 | / | / | / |
| FIM Baseline | / | / | / | / | / | / | 0.36 | 0.051 | <0.0001 |
| CIRS | -1.50 | 0.98 | 0.14 | -0.039 | 0.66 | 0.954 | 0.003 | 0.065 | 0.97 |
| ICU | 31.12 | 25.90 | 0.239 | -5.09 | 7.87 | 0.528 | 1.89 | 1.63 | 0.25 |

6MWT: six-min walk test; CRQ: Chronic Respiratory Questionnaire, FIM: Functional Impairment Measurement; CIRC: cumulative illness rating scale.

Similarly, 13 and eight COVID-19 patients treated at ICU vs. not treated at ICU were used to predict the CRQ at discharge (Table 4). Although the fitting of the model was relative poor (i.e., R^2^=0.40) probably due to the relative small number of patients, the results still suggested that there was no difference in CRQ at discharge between the two subgroups.

The numbers of patients were 26 and 18 in the ICU subgroup and non-ICU subgroup respectively for the model of FIM at discharge. Again, there was no significant difference in FIM scores at discharge between these two subgroups (Table 4).

## Discussion

In this controlled study we show that survivors of COVID-19 pneumonia benefit from our well established in-house rehabilitation program [13, 17] and are improved in physical capacity, disease related quality of life and functional outcome. When compared to patients with common pneumonia included in the same rehabilitation program COVID-19 patients achieve a better outcome in physical capacity and gain a similar disease related disability and quality of life. Regression analyses show that successful rehabilitation outcome is still visible after controlling for confounders such as age, gender, cumulative illness rating scale and baseline values at the start of rehabilitation program. We conclude that in-house rehabilitation is effective and suitable for most COVID-19 patients admitted from hospitals for acute care [18]. Remarkably critical-ill COVID-19 patients treated at ICU could obtain a similar outcome after rehabilitation compared to COVID-19 patients not mechanically ventilated at ICU. Therefore pulmonary rehabilitation is indicated for a fast improvement even after critically-ill Covid-19 infection.

The strength of our study is twofold. It is to our best knowledge the first study on the efficacy of COVID-19 rehabilitation with a control group and considers the clinically relevant improvement as outcome parameter after rehabilitation. Based on our data we are worried by two findings: Firstly, the discrepancy of high physical improvement of COVID-19 patients but their relatively low gains of disease related quality of life (CRQ) compared to the control group. This finding is not explained by a ceiling effect of the CRQ since COVID-19 patients started at a high level of CRQ – as they did for physical performance - but stayed far below the maximum scores at the end of rehabilitation program. Secondly, compared to controls we found a relatively high proportion of 42.9% of COVID-19 patients who failed to improve during rehabilitation. This is again most impressive for disease related quality of life. The reasons why rehabilitation was not effective in such a considerable proportion of patients cannot be fully explained by our data. These patients may develop a chronic disease or rehabilitation program may need adaptions to be optimized for disease specific aspects of COVID-19 rehabilitation. Recent available data indicate that COVID-19 patients after acute care hospitalization still suffer low physical functioning and impaired performances of daily life activities [4]. Whether this is related to persistent abnormal pulmonary function [3] and/or persistent fatigue [5] remains unclear. It is also well known that patients with ARDS which was highly prevalent in our COVID-19 group still show disability after years [2]. If a similar chronic course is confirmed for the COVID-19 pandemic, whole world societies need to consider a strong impact on health care systems in the future.

An interesting finding is that the rehabilitation outcome of our COVID-19 patients is not determined by the need of ICU treatment (Table 4). We however hesitate to interpret this encouraging observation since the study design is not appropriate and sufficiently powered to draw direct conclusions due to small numbers of patients and missing values (especially large number of missing values in CRQ). Future controlled studies are warranted to show if ICU treatment is an important predictor of rehabilitation outcome.

Our study may be limited by a selection bias, since we do not know the clinical course of patients referred to other institutions (e.g. nursery homes) or went to ambulant rehabilitation. Another limitation is CRQ has not been validated for COVID-19 patients, which may also partly explain our results of relative low MCID in CRQ in COVID-19 group compared to the control group. But it is still likely to give an overview of the burden of disease [12] and should be urgently validated in COVID-19 patients considering the exponential growth of the pandemic. Third, the control group was selected from the pulmonary department to provide comparable well established assessment tools. We thus could not fully consider the consequences of COVID-19 associated vascular complications in our study.

## Data Availability

The database generated during and/or analysed during the current study are available from
the corresponding author on reasonable request.

## Disclosure

The authors declare no conflict of interests.

## Notes

### Competing Interest Statement

The authors have declared no competing interest.

### Funding Statement

There is no funding for this study

### Author Declarations

All Patients gave written general consent for using their data for research purpose. The patients data were anonymised before analysis. This study was approved by the scientific committee of Academy Barmelweid.

